# Alcov2: a National Questionnaire Survey for Understanding the Transmission of SARS-CoV-2 in French Households during First Lockdown

**DOI:** 10.64898/2026.02.23.26344954

**Authors:** Amaury Lambert, Anna Bonnet, Pierre Clavier, Pierre Biousse, Lucas Clavières, Sophie Brouillet, Sylvie Chachay, Marie Jauffret-Roustide, Sonia Lewycka, Nicolas Chesneau, Grégory Nuel

## Abstract

We describe a fast, noninvasive, low-cost survey method designed to understand the mode of transmission of an emerging pathogen. It is inspired from the standard household prevalence survey consisting in sampling households and counting the total number of people infected in each household, but refines it with the aim of improving diagnosis and estimating more parameters of the model of intra-household transmission.

The survey was carried out in May-June 2020, during part of the first national French lockdown and received responses from more than 6,000 households involving a total of 20,000 people. We explain how we conceived the questionnaire, how we disseminated it, to the public through an open website hosted by CNRS, marketed through media and social media, and to a socially representative panel hosted by two survey institutes (BVA, Bilendi). We used the data obtained from the representative panel to correct for sampling biases in the CNRS survey using a classical raking procedure.

Our results indicate that raking correctly canceled statistical biases between the two populations. We obtain the empirical distribution in households of the number and nature of symptoms.

The main factors affecting the presence of symptoms are age, gender, body mass index (BMI), household size, but not necessarily in the expected direction.

Our study shows that combining self-reporting and representative surveys allows investigators to obtain information on prevalence and household transmission mechanisms on emerging diseases at low cost.

## 1 Introduction

In order to fight and curb an epidemic like the COVID-19 pandemic, health professionals, epidemiologists and policy makers need to gather data to understand the specificities of the infection process and to assess the progress of the epidemic.

By investigating the number of infected individuals in any particular collective social unit (e.g., household, firm, school, university, sport club, choir, etc.), large-scale, randomized surveys can estimate the prevalence of a disease in a country (Jauffret-Roustide et al., 2009; Vogl et al., 2021), especially when such studies are aggregated across units (of the same type or of different types). More importantly, such surveys can help to understand the modes of transmission of the disease by contrasting the results obtained in the presence vs absence of masks, in crowded vs spacious places, indoor vs outdoor places, ventilated vs not ventilated rooms, etc., sometimes using mathematical models linking the number of cases with the parameters of the transmission process (Harris, 2021; Curmei et al., 2021; Kuwelker et al., 2021; Bi et al., 2021). These estimates can in turn help inform health agencies and also record current trends to be contrasted with future data, in order to assess possible changes in the epidemic process due, for example, to pathogen evolution, vaccination, seasonality, or shifts in social behavior.

In addition to collecting data on say, the sociodemographic characteristics, lifestyle, behavior and health condition of the respondents, the survey procedure has to collect information on their disease status (infected vs non-infected), coping with a possibly high degree of uncertainty. There is a whole gradient of uncertainty, but we can distinguish three main types of situations.

In the optimal case where the disease has unambiguous, obligatory symptoms, the survey merely amounts to counting individuals who have presented these symptoms. Second, if the most frequent symptoms of the disease of interest are shared with other diseases, but the disease of interest is sufficiently well understood that diagnosis tests detecting the presence of antigens of the pathogen or their associated antibodies, are reliable and easy to acquire, the survey can consist of applying the test to biological samples taken from randomly selected social units (e.g., households). We note that this procedure may be cumbersome both in terms of administrative/ethical guidelines and of financial costs (González López-Valcárcel and Vallejo-Torres, 2021). In the third case where the disease not only has ambiguous symptoms, but is also emergent like COVID-19 was in 2020, the task is made even more difficult for several obvious reasons, including the fact that tests may have low reliability and/or be too expensive or in short supply. This may have direct consequences on both the number of surveys that are conducted and their reliability, with the two factors interacting negatively (Cai et al., 2023).

The methodology we propose here applies to any emerging disease with possibly ambiguous symptoms (here, COVID-19). It consists in distributing questionnaires to social units (here, households) asking about the presence of symptoms from an exhaustive list of symptoms potentially related with the disease, along with other personal data like health condition (e.g., chronic disease) and/or social characteristics (e.g., number of rooms in household).

The rough data thus collected at low cost can rapidly inform health agencies of the state of the epidemic. The deeper, longer-term goal of such a study is two-fold: 1) to develop an algorithm mapping personal data (a list of symptoms and health/social features) to a *diagnosis score* between 0 and 1 equal to the probability of being infected by the disease of interest conditional on personal data, and 2) to design a *probabilistic model of transmission*, ideally letting the transmission rates depend on health/social features of donor and receiver and taking into account hidden variables like the number of non-symptomatics or non-infectious individuals, and use it to estimate these rates.

The algorithm developed in the first task can serve as a low-cost diagnosis test while biological diagnosis tests are still under testing phase or in short supply. The estimates (of rates parameterizing the infection process) obtained in the second task can inform epidemiologists and health policy makers of various key quantities (e.g., frequency of asymptomatics, distribution of time between infection and first symptoms) improving the knowledge of the epidemic dynamics of such emerging pathogens.

To perform the first task (diagnosis scoring), we can leverage on three sources of information: 1) previous knowledge on correlation between symptoms, health features and disease susceptibility; 2) information coming from the respondents of the survey who had been tested; 3) knowledge of the number of other household members with characteristic symptoms (e.g., loss of sense of smell or anosmia in COVID-19).

The second task (parameter estimation) typically relies on stochastic models like the Reed-Frost model (Reed and Frost, 1952; Abbey, 1952; Bailey, 1975; Picard and Lefevre, 1990), parameterized by the daily rate of transmission and based on one summary statistic like the total number of household members infected after *x* days. We add here an extra layer of information by adding to the questionnaire a section about the date of appearance of first symptoms of each household member. This extra information will help us infer with more confidence the chain of transmission and to estimate with more accuracy the daily transmission rates and the rates of non-symptomatics and/or non-infectious (Tsang et al., 2023).

We will describe elsewhere the detail of how we perform these two tasks. In the present paper, we expose the methodological process of data collection according to how we have proceeded to conceive and distribute the questionnaire, we explain how we have corrected the data for possible sampling biases and we give a broad overview of the data collected, as outlined below.

The next section (Section 2) deals with material and methods: context about the first French national lockdown in spring 2020; details about the conception of the questionnaire and its distribution through two channels: an open online questionnaire and a representative survey sample; technical aspects of the statistical procedure, called ‘raking’, aimed at correcting the former dataset via the latter.

Section 3 displays our main results: effect of media campaigns on questionnaire fillings, effect of raking on the dataset obtained from the online questionnaire, description of data (after raking): distribution of the number and nature of symptoms, differences between the two datasets, influence of social factors on transmission (body mass index, age, gender, household size).

In Section 4, we discuss our results: benefits and limitations of the procedure, possible improvements, knowledge gained about household transmission of SARS-CoV-2.

## 2 Material and Methods

### 2.1 The first French national lockdown

Following the exponential increase of COVID-19 cases and hospitalizations in France and neighboring countries in February and March 2020, a national lockdown was declared in France starting March 17 (March 9 in Italy, March 15 in Spain). The French lockdown ended on May 11, after two renewals on March 27 and April 13. During this period, only ‘essential workers’ (healthcare, police, public transports, food stores, etc.) were allowed to travel outside their home. For all other people living in France, the only permitted reasons for traveling were food shopping, medical appointments, family emergencies (including shared child custody), physical exercise near home (Le Monde, 2020).

The lockdown period was a life-size experiment in which virtually all French households were subjected to similar living conditions. This homogeneity was ideal to infer the general rules of within-household virus transmission. Of course, living conditions differed in some aspects, but most of these aspects were known, and we addressed them in the questionnaire: house size, number of household members, their age and gender, presence of essential workers, medical condition, etc.

### 2.2 Data collection

#### 2.2.1 Questionnaire development and content

The questionnaire (see logo Figure 1) was distributed through two channels: an open online questionnaire thanks to the help of public research facilities, hereafter called the ‘CNRS’ survey (acronym of the French National Center for Scientific Research), and a representative survey, thanks to the help of survey institutes BVA and Bilendi, hereafter called ‘BVA-Bilendi’ survey. The integral content of the questionnaire is reproduced in Appendix A. The labels of questions below refer to the labeling in Appendix. Answers coded as ‘DK’ mean ‘Don’t Know’ (‘*Ne se prononce pas*’ or ‘*Ne sait pas*’ in French).

**Figure 1:**
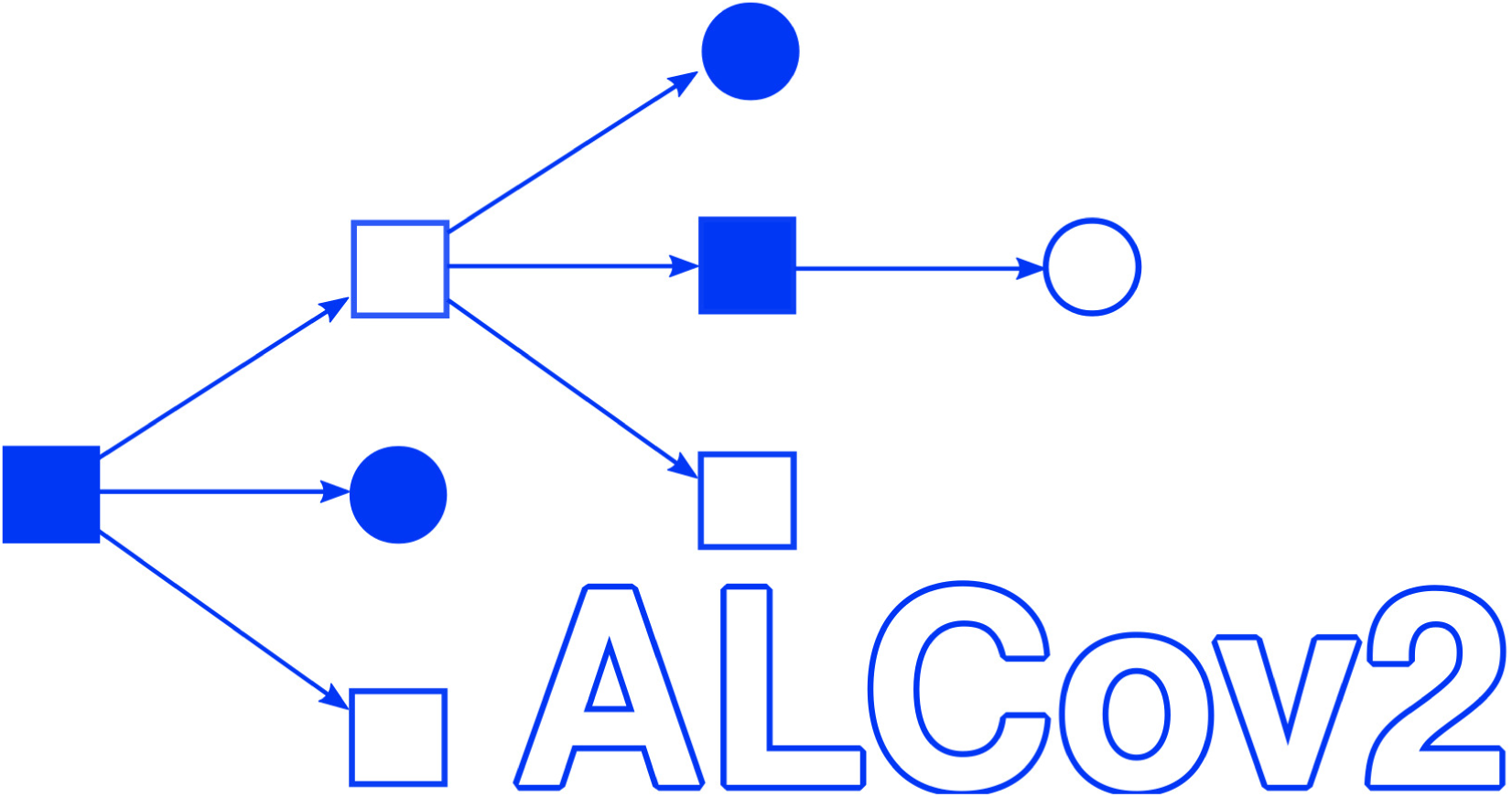
Logo of the questionnaire survey.

The questionnaire aims to collect all data available on (1) nature of symptoms (if any) of all household members, in order to infer disease status of each person in the absence of test; (2) dates of infections and level of exposure during lockdown, in order to infer the dynamics of infections from outside and within the household (who-infected-who); (3) risk factors, to help infer both types of information; (4) sociodemographic information (different in CNRS and BVA-Bilendi surveys), to detect geographical differences and to correct for sampling biases. At the onset of the French national lockdown, people were allowed to travel once to choose and stay confined in another place than their usual home. We refer to this place as the ‘confinement residence place’. Since we were mainly interested in the dynamics of the epidemic within households during lockdown, the questions only concern the configuration of the household in the confinement residence place.

Because we sought to study the process of introduction of the virus into the household and especially propagation of the virus within the household, we were not interested in households with no sick member or hosting only a single member. Therefore, the questionnaire starts with a warning announcement, prescribing to complete the survey only if the following two conditions are fulfilled:

1. At least one person living in the confinement residence place has recently^1^ experienced symptoms that could be related to Covid-19, even if uncertain that they are really due to Covid-19;
2. At least two people live in the confinement residence place.

Because collecting all this information relies on the good will of questionnaire respondents, the conception of the questionnaire was subject to a trade-off between maximizing the amount of information collected and minimizing the duration of questionnaire filling, by moderating the number of questions and reducing their ambiguity as much as possible.

Questions have been formulated so as to limit biases of social desirability, i.e., the tendency to report an answer thought to be more socially acceptable to interviewers (Crowne and Marlowe, 1960).

**Questions regarding the nature of symptoms (1).** In order to infer which household members have been infected, the questionnaire asked (Q-II.10) the respondent the following question concerning each household member: “Since mid-February 2020, has this person experienced one or more symptoms, even mild, possibly related to Covid-19? (for example: fever, cough, headache, sore throat, fatigue, diarrhea, chills, nausea, loss of taste or smell, shortness of breath, chest pain, etc.)”.

After receiving reports from respondents having experienced characteristic symptoms as early as November or December 2019, we removed from the previous question the mention to ‘mid-February’ to lift the restriction to symptoms declared only after this date.

If the respondent answered ‘Yes’ (other possible answers: No/DK), we called this household member a ‘case’, even if we do not know with certainty whether these symptoms are due to COVID-19 or not.

The questionnaire then repeats the same set of questions for each ‘case’ successively.

First, for each case, the respondent is asked to answer Yes/No/DK to each item in the extensive list of possible symptoms (Q-IV.1). The list of symptoms was elaborated with the help of clinicians and was finalized as fever, chills, unusual fatigue, headaches, loss of sense of smell or taste, muscle aches, cough, shortness of breath, chest pain or tightness, abdominal pain, nausea, diarrhea, nasal congestion, runny nose, conjunctivitis, skin rashes. To avoid ambiguity, we specified: ‘fever (*>*38°C)’, ‘diarrhea (three soft stools in a row)’, and ‘runny nose (except spring allergy)’.

We added complementary questions to obtain information on the severity of symptoms (medical attention Q-IV.2 and/or hospitalization Q-IV.6) and the existence of external diagnoses (by physician Q-IV.3 and/or test Q-IV.4-5). To account for uncertainty in the diagnosis, we proposed the following multiple answers: Covid-19 unlikely/possible/likely/near-certain, moderate form/near-certain, severe form/DK.

We resolved potential doubts on the terms ‘positive’ and ‘negative’ for the result of the test by giving the choices: Positive (infection)/Negative (no infection found)/DK. To avoid missing any potential input from the respondent, one last question concerned the respondent’s opinion on the disease status of each person (Q-IV.7).

**Questions regarding dates of infections and level of exposure (2).** Since the dates of infection are unknown, we asked for each case the date of symptom onset (Q-III). Because we have a better memory of durations than of dates, we encoded these data in the form of *t*_1_, *t*_2_ − *t*_1_, *t*_3_ − *t*_2_…, where *t_i_* is the date of symptom onset of the *i*-th household member with symptoms (in chronological order of first symptoms).

To avoid any ambiguity when asking to count people living in the household, we asked (Q-I.2): “How many people live in your confinement residence place (including yourself and children even in shared custody)?”, resolving the ambiguity between ‘usual residence’ and ‘confinement residence’ and the question of children in shared custody.

We also needed to determine how intensely household members interact on a daily basis. To do this, we collected data on home size and on the time shared within the household by its members. A common way to score the size of a home is through its number of bedrooms (Q-I.3). As for time spent at home, to avoid any ambiguity, we asked accurate questions: absence for at least 3 consecutive days and nights since the beginning of lockdown Q-II.3, absence at least 3 days a week due to work or activity Q-II.4. In order to score the possibility of infections from outside, we added a question concerning persons having answered ‘Yes’ to Q-II.4 and asking whether the activity led to contacts with the public or not (Q-II.5), distinguishing between ‘hospital or medical facility’ or ‘other’ (sales, public transport, police, etc.).

**Questions regarding risk factors (3).** Here again, we requested the help of clinicians to elaborate the list of potential risk factors. We collected information on the following factors: age, sex, body mass index (BMI), BCG vaccine, pregnancy, smoking, chronic disease. The questions about risk factors were repeated for all household members, not only persons with symptoms, because the absence of symptoms is as informative as the presence of symptoms in terms of interaction with risk factors.

We only let two options for sex, that we called ‘biological sex’, because we are interested in the effect of sex, rather than of gender, in the experience of the disease. We did not ask for BMI directly, but only asked for the height and weight of each person in the household older than 2 years (Q-II.2). For BCG, we reminded the respondent that the vaccine is injected into the upper left arm and sometimes leaves a small scar there (Q-II.6). Pregnancy was only considered from 3 months’ duration (Q-II.7). Smoking was characterized as ‘smoking/vaping at least once a day’ and divided into 5 categories: Yes, now/Yes, in the last 3 years/Yes, in the last 10 years/Never/Other. Chronic diseases were divided into diabetes, respiratory disease, heart or vascular disease, kidney disease, liver disease, immunosuppressive disease or treatment, other (Q-II.8).

**Questions on sociodemographic data (4).** All respondents were asked the department^2^ of their confinement residence place (Q-I.1).

Some additional sociodemographic data were collected in the BVA-Bilendi survey for quota adjustment, and not in the CNRS survey. Members of the BVA-Bilendi panel are used to answering these types of question, whereas voluntary respondents to the CNRS survey could have judged these questions too intrusive and thereby been deterred to respond. In addition, approval by data protection regulation authorities would have been longer and more rigorous if we had added these questions to the CNRS survey. In turn, information collected in the BVA-Bilendi survey is used to correct sampling biases in the CNRS survey (see below).

The questions specific to the BVA-Bilendi survey are: zip code and name of municipality of the usual residence place, socio-professional category of respondent/of household reference person (if different). See Appendix A for details.

#### 2.2.2 Implementation

Like many big universities, Sorbonne Université hosts a facility (“*Pôle Intégration et Développements*”) employing engineers trained to code surveys for research purposes. Unfortunately, at the time of our request (early April), all the technical staff of Sorbonne Université was forced to take a week’s vacation. We then turned to the GRICAD facility (“*Grenoble Alpes Recherche – Infrastructure de Calcul Intensif et de Données*”), a support unit hosted by the Math Institute (INSMI) of CNRS and Université Grenoble-Alpes. GRICAD offered to host and code the CNRS version of the questionnaire using the online survey tool *limesurvey*. It took us (AL, SB, SC) two weeks working off hours to finalize, code, test, and debug the survey before finally launching it on April 27. Further debugging was performed along the way thanks to feedback from users via hotline email address.

In parallel, the BVA-Bilendi survey was handled by the technical staff of BVA and Bilendi, teamed up with the first author, and launched on May 6.

#### 2.2.3 Data protection and scientific board

In the European Union, personal data protection laws are gathered under the EU-GDPR act (General Data Protection Regulation) and enforced in France by the CNIL (“*Commission nationale de l’informatique et des libertés*”). Every firm or university collecting personal data must be endowed with a DPD (Data Protection Delegate) in charge, for each data collection procedure, of ensuring that legal provisions are respected and of making a declaration of compliance to the CNIL.

The Alcov2 project relates to ‘research that does not involve the human person’ and falls under CNIL’s MR-004 reference methodology. Hosting our data in a certified health data center was not compulsory because the data have not been collected during ‘prevention, diagnosis, care or social and medico-social monitoring activities’. Before filling out the questionnaire, respondents give their agreement that their data will be used for scientific research and are informed that their data will not be used for commercial purposes and that they will be processed pseudonymously and aggregated.

The questionnaire was anonymous to avoid deterring the respondents from filling it in. However, by French legislation, respondents have the right to access and correct their personal data. This could be done by providing a location (‘*département*’) along with date and time when the survey was filled, although this pseudonymization is not very convenient and can be ambiguous, so not safe (alternative solutions should be preferred).

The data were stored on secured servers, first by GRICAD (server hosting the survey), then by the firm Ekimetrics (under contract with Sorbonne Université for processing and analysis of the data) and finally on the servers of the institute of computing and data sciences (ISCD) of Sorbonne Université.

To assist survey designers and analysts, a scientific board was named that represented several fields of research relevant to the Alcov2 study: Jean-Christophe Thalabard (statistics, endocrinology, Cochin Hospital), Vincent Maréchal (virology, Sorbonne Université), Odile Launay (infectiology, Cochin Hospital), Marie Jauffret-Roustide (sociology, health, INSERM), Isabelle Hilali (data science, health, datacraft).

#### 2.2.4 Questionnaire dissemination

As previously emphasized, the survey was distributed through two independent channels. The goal of this double dissemination was to obtain both a massive dataset and a representative one. The CNRS survey has been free access from late April to late June 2020. The dissemination of this survey has been promoted through an intensive media campaign (press, television, radio, social media), see Section 3.2. The BVA-Bilendi survey was submitted by the BVA survey institute at roughly the same dates to a representative panel of the French population of 10,000 people, hosted by the Bilendi survey institute.

The preamble of the questionnaire specified which households were eligible to fill it – at least two persons in the household, at least one with Covid-related symptoms, see Section 2.2.1. The respondents to the BVA-Bilendi questionnaire represented only a fraction *p* of the Bilendi panel, roughly equal to the prevalence of Covid-19 in France in spring 2020 (more rigorously, the expectation of *p* is the probability of living in a household having experienced Covid-related symptoms, comprising at least two people, thinned by the willingness to respond to this kind of survey – members of the panel are asked to complete surveys on a regular basis, approximately twice a month, and are not obliged to complete all of them). The advantage of the BVA-Bilendi survey is two-fold: first, it gave us a guarantee of getting at least ≈ 10,000 × *p* completed questionnaires; second, it contains other variables that can be used to correct the CNRS dataset for sampling biases, see Section 2.3.2.

The responses to the CNRS survey depended on two types of impulses: media advertizing and word-of-mouth/social media transmission. The contribution of both dynamics to the spread of the information is modeled in Section 2.3.4.

One difficulty with word-of-mouth/social media transmission is that the information on the survey can remain circulating in the same social circles for a long time, as in so-called snowball samplings. Léon et al. (2016) It is hard to know if the information really manages to reach social circles far away from its origin (here, Alcov2 researchers and their friends). Another difficulty, specific to surveys concerning only people having had a specific experience (here Covid-19), is that people willing to spread the information most frequently target people they know have had this experience, which only represents a fraction *p* of their contacts, instead of sending the information to all their contacts. This results in chains of transmission rapidly going extinct, because this thinning by *p* often results in a small number of target contacts.

Interviews in the media are expected to trigger responses to the survey by a relatively diverse sample. The size of this sample greatly depends on whether the journalists transmitted the URL of the questionnaire.

### 2.3 Statistical Methods

#### 2.3.1 Handling outliers

We noticed that our dataset contained outliers, which are values that we considered either inconsistent or that we could not use. More precisely, we encountered the following types of outliers:

- Given the length of the questionnaire which is substantial, if completion times are less than 3 minutes, we considered that it was likely to contain errors. Similarly, we decided to consider an outlier an individual whose answering time was too long, typically greater than 35 minutes.
- For the house size, we considered outliers houses with zero bedroom.
- Regarding the body mass index (BMI), for which values above 40 correspond to morbid obesity, values above 60 were treated as outliers and set to NA.
- Households with more than two outliers in BMI were considered outliers.
- BMI for individuals younger than 18 years is considered irrelevant and therefore set to NA.

The flow chart of households is shown in Table 1. The modified entries are detailed in Table 2.

**Table 1:**
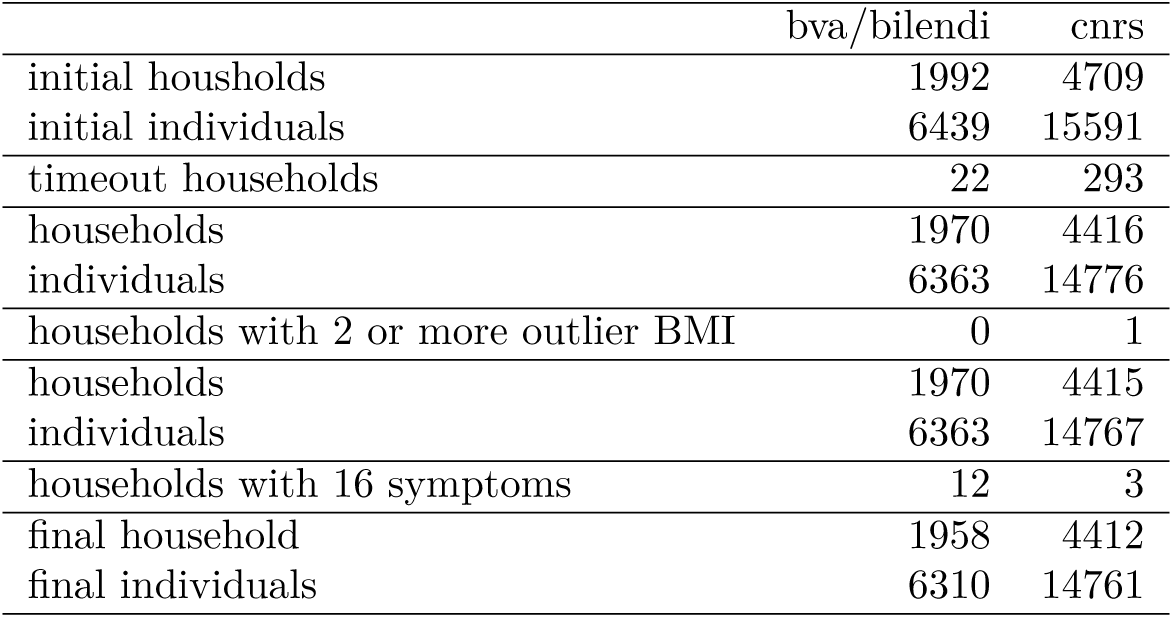
Households flow chart.

**Table 2:** Modified entries.

|  | bva | cnrs |
| --- | --- | --- |
| households with 0 room set to 1 room | 0 | 19 |
| outliers BMI set to NA | 18 | 10 |
| BMI for age <18 set to NA | 1145 | 3207 |

Since several features concern the whole household, when we found an outlier value for one individual, we decided to remove their household for the dataset. Eventually, 331 households composed of 957 individuals were removed from the survey.

#### 2.3.2 Statistical calibration: raking procedure

**Raking.** Here we describe how we performed the raking (Deming and Stephan, 1940; Deville et al., 1993). Given a population *U* of size *N*, a sample *S* of size *n* is randomly selected, where individual *k* is chosen with probability 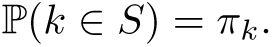 Each variable is observed only in the sample *S* and not in the total population. Our goal is to estimate for a reference binary variable *Y* (e.g. 1 if headache, 0 if not) the mean 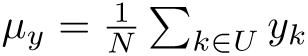 or the sum 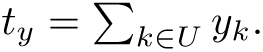

A typical estimator called *Horvitz-Thomson estimator* (Horvitz and Thompson, 1952) of the sum *t_y_* is the weighted sum on the sample *S*:

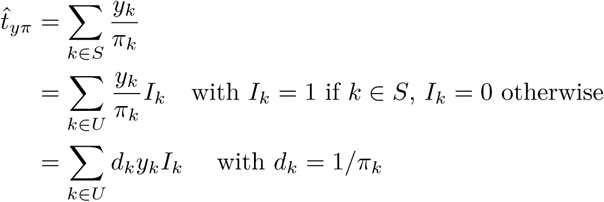

A natural choice for the weights *d_k_* is to set *d_k_* = *N/n* for each *k*. However, these weights must be adjusted for biased sampling. For example, if we study a feature which is impacted by gender, we will want our sample to contain men and women in the same proportions as the whole population. More generally, we want to account for the effect of *p* variables 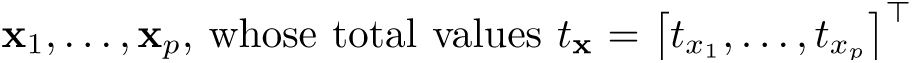 are known in the representative population (here the BVA-Bilendi dataset). Then we aim at finding new weights *w_k_*, close to the previous weights *d_k_*, which verify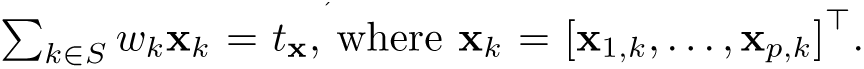 We define the vector 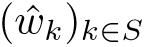 as the minimizer of

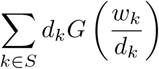

under the constraint 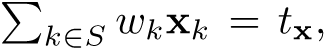 where *G* is a convex cost function such that *G*(1) = 0. One way to ensure that we obtain interpretable positive weights is to choose *G*(*r*) = *r* log(*r*) − *r* + 1 (called the raking ratio method). We use a Newton-Raphson algorithm to perform the minimization.

**Statistics selection for raking.** We perform raking by correcting only on general epidemiological statistics and risk factors. Namely, we correct for the frequency of each of the following features:

- Household location: Paris, south of France, other;
- Individuals in the household: 2, 3, 4, 5, 6 or more;
- Rooms in the household: 1, 2, 3, 4, 5, 6 or more;
- Individuals in each of the following age bins: 0-18, 18-25, 25-40, 40-55, 55-65, 75 and more;
- Individuals in each of the following BMI bins: 0-18, 18-25, 25-30, 30-35, 40 and more;
- Individuals in each of the following bins counting the number of chronic diseases: 0, 1, 2 or more;
- Individuals moving outside the household, moving for medical reasons, moving for other reasons;
- Number of pregnant women;
- Number of males;
- Number of smokers.

#### 2.3.3 Statistical inference

We performed a multivariate logistic regression where the observations are binary (presence/absence of symptoms) and the covariates are the different features: BMI, age, number of rooms, number of people in the household, gender and location. In addition, we used standard statistical approaches to assess the features that are significantly associated with the presence of symptoms, such as likelihood ratio tests thanks to the R package car. All analyses, performed with R, account for the weights obtained with the raking procedure described in Section 2.3.2.

#### 2.3.4 Time series analysis

To study the effect of the information campaign in the media on the number of people who answered the questionnaire, we represent the average number of respondents around the time of media interventions. More specifically, if *x* = (*x*_1_ *,…, x_m_*) are the dates of media interventions and *y* = (*y*_1_ *,…, y_n_*) are the dates of survey fillings, we display the graph of the function F defined as:

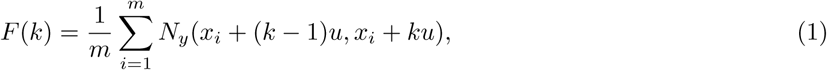

where *N_y_* (*I*) is the number of occurrences of survey fillings *y* on the interval *I* and *u* is the length of the intervals considered (in practice, we choose *u* = 1 day). For example, *N_y_* (*x_i_, x_i_* + 1) is the number of people who answered the survey within one day after the *i*-th media intervention, and *F* (1) is the average number of survey responses within one day of all media interventions. The latter value should be high if the media campaign had a positive impact on the number of answers, although, of course, it does not by itself prove any causality.

To investigate the efficiency of the media campaign in specific regions, we determine if there are geographical differences in the distribution of respondents after the information campaign. We use a *k*-means algorithm to identify departments with the same filling profiles, where these profiles are entirely determined by the function *F* defined in Eq (1), normalized by the number of respondents in each department. The number of classes that we give as input to the algorithm has been set to 2, since the preliminary analysis revealed that it provides groups with very separate and interpretable profiles.

## 3 Results

### 3.1 CNRS and BVA-Bilendi datasets

The CNRS dataset contained the answers from 4,709 households and the BVA-Bilendi dataset 1992 households with at least one member having any COVID-19 symptoms, gathering information on about 15591 and 6439 people, respectively.

### 3.2 Results: media campaign and number of fillings

#### 3.2.1 National results

The survey was opened 8 days before the start of the media campaign that lasted 3 days. When considering the distribution of answers at a national scale (Figure 2), we observe a first peak, corresponding to the opening of the survey, then a decrease, followed by another peak with a sharp maximum centered at day 0 that indicates an increased average number of fillings immediately after media interventions. This observation suggests the efficiency of the media campaign. Note that the second peak starts before 0, which can be explained by the fact that we consider the number of responses averaged over all media interventions and these interventions happened in a very short time (8 interventions in 3 days).

**Figure 2:**
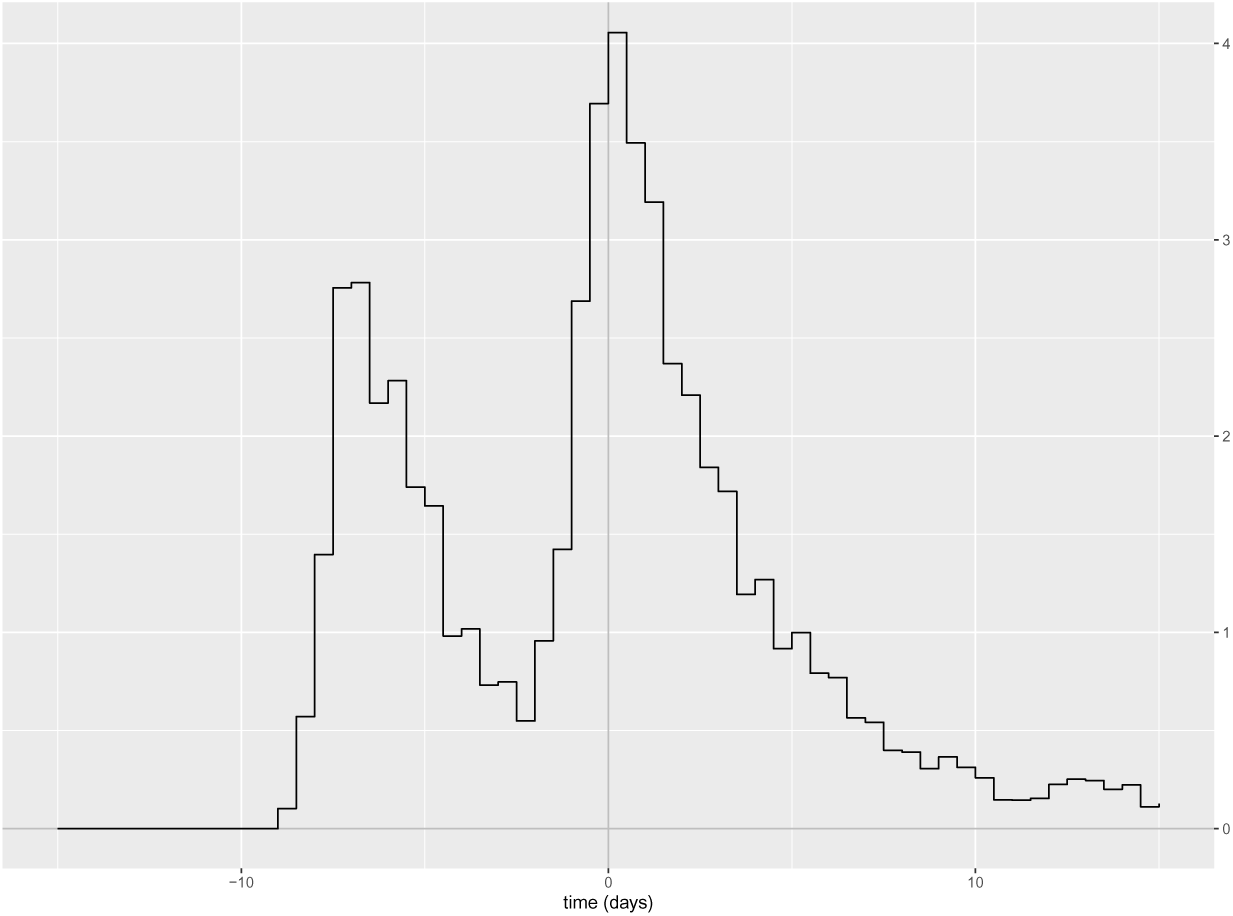
Distribution of the CNRS survey responses averaged around all media interventions, which correspond to time 0. Vertical scale unit: 100 households per day.

#### 3.2.2 Geographical differences

We provide the distribution of the number of answers in each department (see footnote 2 page 5) as input to a *k*-means algorithm to investigate whether this distribution would vary according to a geographical criterion. The *k*-means algorithm inferred the presence of two clusters of households exhibiting two different profiles with respect to the response following media interventions (Figure 3). In the first group, a large number of responses were given at the opening of the survey, followed by a second peak after the start of the media campaign. In the second group, very few responses were registered when the survey opened; however, many people answered after the beginning of the media diffusion: this suggests that in these regions, people have become aware of the survey only when it was spread by the media. Interestingly, most of the large cities are part of the first group.

**Figure 3:**
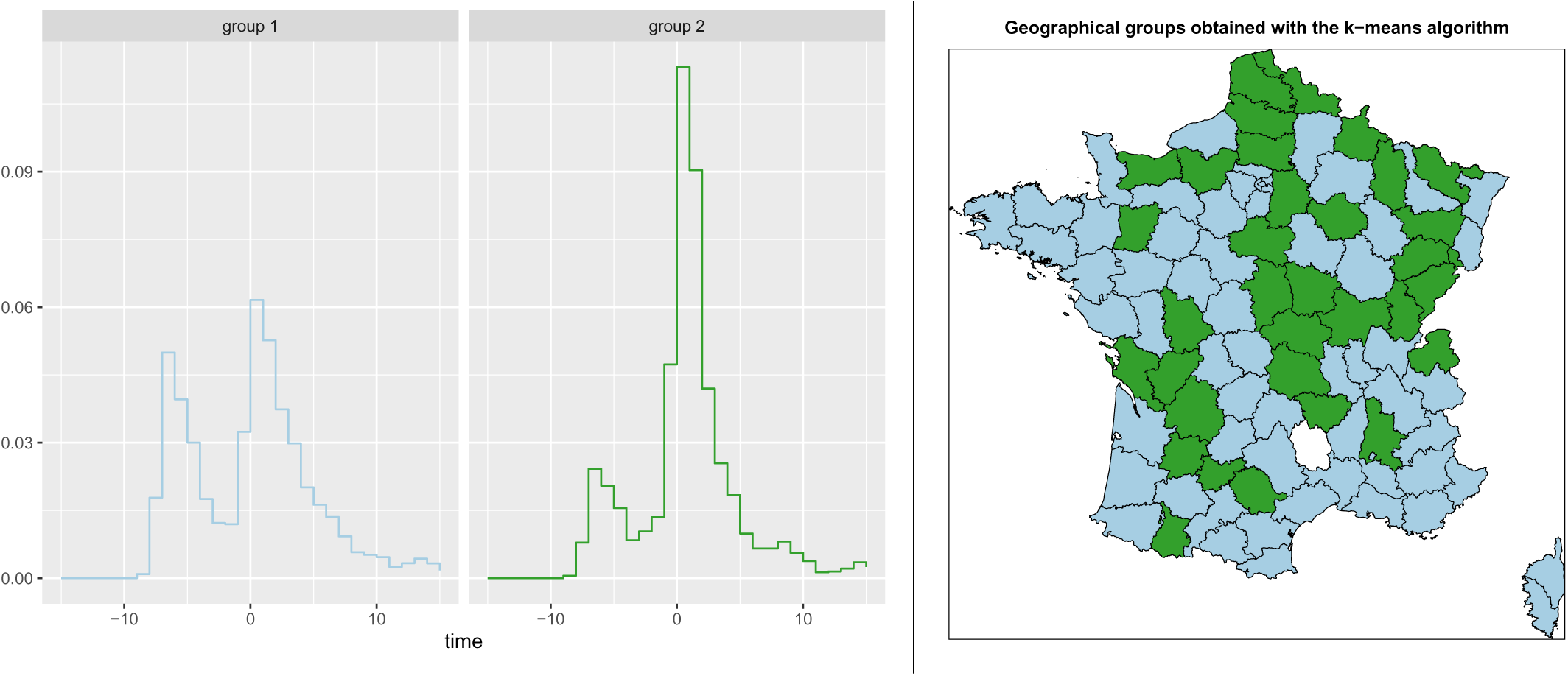
Left: Average distribution of the CNRS survey responses around media interventions in each of the two groups obtained with the *k*-means algorithm (the distributions have been normalized by the total number of fillings in each group). Right: Geographical distribution of the two groups.

We can then assume that the media campaign significantly increased the number of respondents to the survey. Moreover, it allowed us to access a sample with more geographical diversity, in particular, the departments of the aforementioned second group.

### 3.3 Results of the raking

Figure 4 displays the distribution of weights associated with the CNRS households. The values are between 0.08 and 5.75, most of them between 0.2 and 2 with an average weight equal to 0.79, which shows that the CNRS dataset will be underweighted overall. Let us focus on the households that have been associated with extreme (high and low) weights by the raking procedure (see Table 3). First, note that the smallest coefficients obtained with the raking procedure correspond to weights that have been divided by up to 12, while the large coefficients correspond to weights that have been multiplied by around 5. This suggests that the major effect of the raking procedure is to decrease the influence of some households from the CNRS dataset, but without balancing with very high weights for others. The high-weighted households contain categories of people that are underrepresented in the CNRS dataset: obese adults (BMI greater than 30), adults over 65 and people living outside Paris region. Regarding the low-weighted households, we note that they are all from the Paris region, they contain neither obese nor individuals over 65 and finally they all have a large number of rooms.

**Figure 4:**
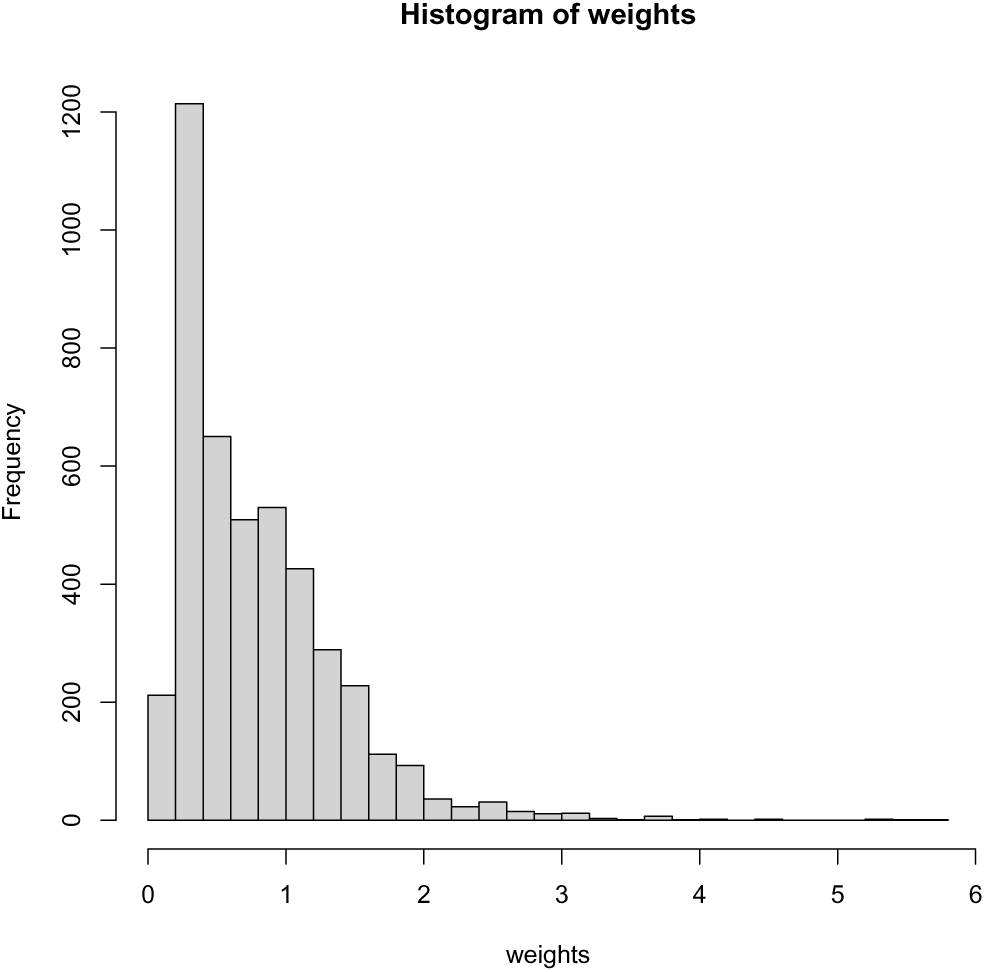
Histogram of the 4412 CNRS households weights after raking: maximum weight 5.75, minimum weight 0.0806, mean weight 0.79.

**Table 3:**
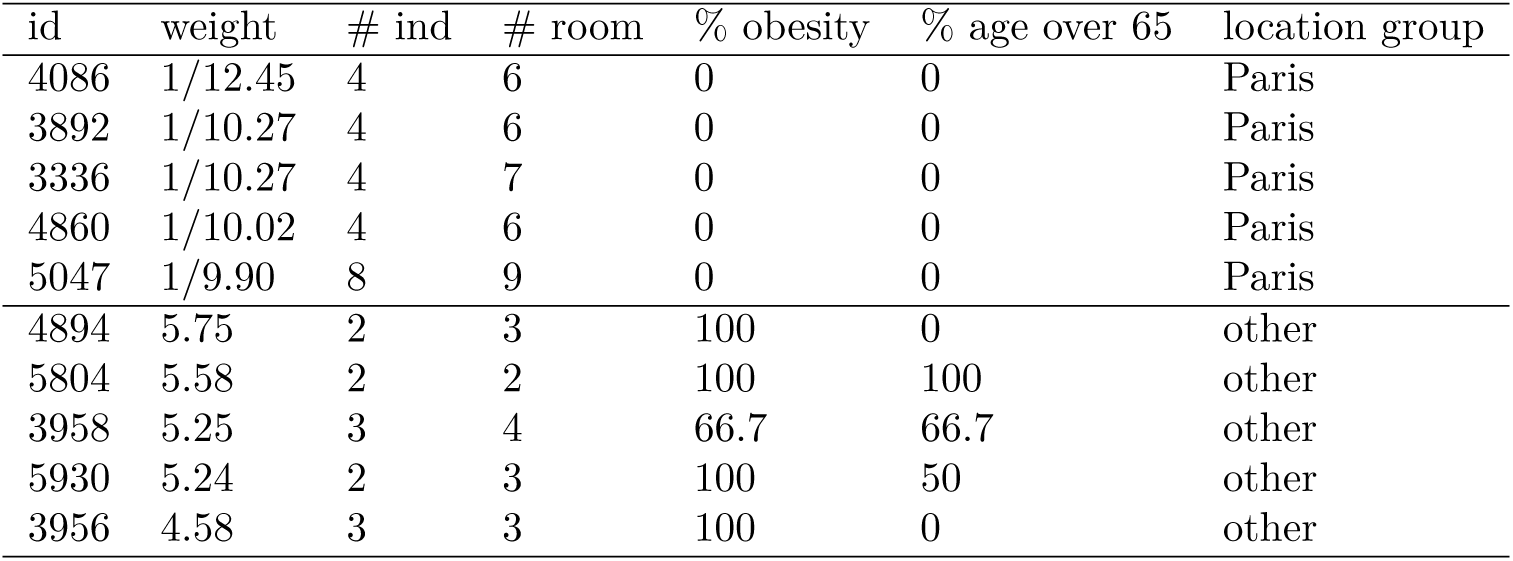
The ten most extreme household weights and some household statistics: ID of the household, number of individuals, number of rooms, percentage of obese adults in the household (where obesity is defined as a BMI greater than 30), percentage of people over 65 in the household, location and location group (divided in Paris, South and other).

**Table 4:** p-values associated with the likelihood ratio tests for each feature in the logistic regression. The number of stars indicates the significance of the test: *** (p <0.001), ** (0.001<p<0.01), * (0.01<p<0.05).

| Feature | p-value likelihood ratio test |
| --- | --- |
| Age | $< 2.2 \times 10^{-16}$ *** |
| BMI | $1.8 \times 10^{-5}$ *** |
| Number of rooms | 0.0018 ** |
| Number of people | $< 2.2 \times 10^{-16}$ *** |
| Gender | $< 2.2 \times 10^{-16}$ *** |
| Location | 0.00092 *** |

Figures 5 and 6 illustrate the performance of the raking. More precisely, Figure 5 shows that the original CNRS dataset contains significantly less people with overweight (BMI greater than 30) and more people living in Paris than the BVA-Bilendi dataset. The age distributions were also quite different: in particular, the original CNRS dataset was composed of more children and less people over 65 years of age. All these effects have been taken into account in the adjustment procedure, which efficiency is confirmed by the almost identical distributions between the BVA-Bilendi and the CNRS dataset after raking.

**Figure 5:**
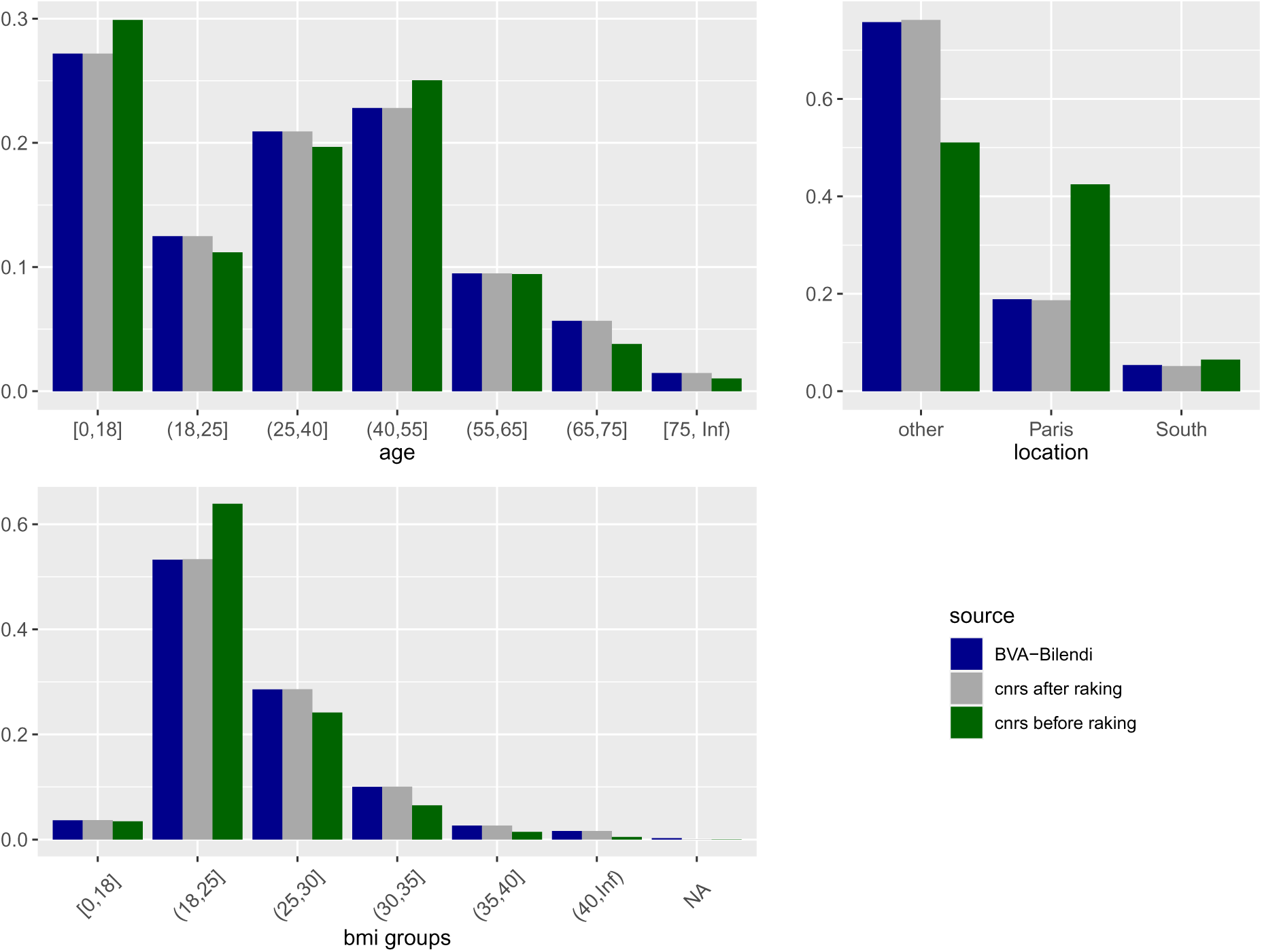
Distribution of BMIs (of adults only), ages and localization (divided in three groups: Paris region, South of France, other) in the BVA-Bilendi dataset and the CNRS dataset, before and after raking.

**Figure 6:**
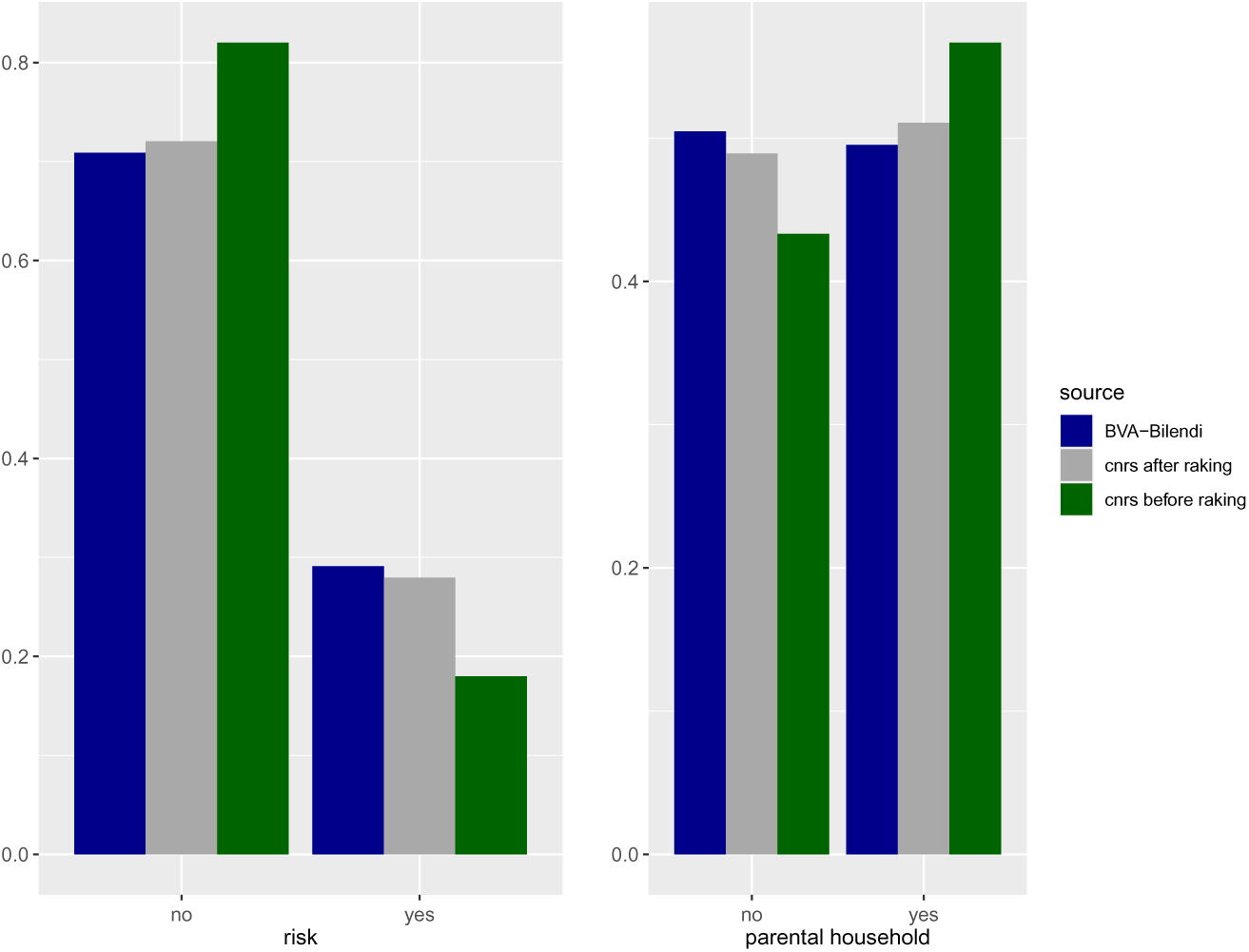
Left: Proportion of households with at least one individual with higher risk factor (age greater than 75 and/or BMI greater than 30). Right: Proportion of ‘parental households’, which are composed of two adults (between 25 and 65) and at least one child (less than 18). All results are given in the BVA-Bilendi dataset and the CNRS dataset, before and after raking.

Interestingly, the raking procedure also allows homogenizing the datasets according to criteria that were not directly accounted for when computing the raking weights. For example, we note that the CNRS dataset contains a large number of households composed of two adults (aged between 25 and 65 years) and one or several children. Figure 6 shows that this difference between the BVA-Bilendi and CNRS datasets has been smoothed by the raking. Similarly, we defined a category of people at risk (age greater than 75 and/or BMI greater than 30), which are more numerous in the BVA-Bilendi dataset than in the CNRS dataset: this difference almost disappears after raking.

### 3.4 Variables on which we did not perform an adjustment

Some variables such as, for example, the proportion of individuals with symptoms, were excluded from the raking procedure. Indeed, there are objective reasons why some proportions may vary from one dataset to another, due to the different ways of spreading the questionnaire. All participants in the BVA-Bilendi dataset were asked to complete the survey, while the CNRS respondents chose to respond, which could result in differences in the proportion of people with symptoms or in the personal opinions on whether they were infected or not (see Figure 7). Interestingly, more people in the CNRS dataset believe they are likely to have suffered from Covid-19 (Figure 7, bottom panel), and they also show more symptoms (Figure 7, top panel) than the BVA-Bilendi respondents.

**Figure 7:**
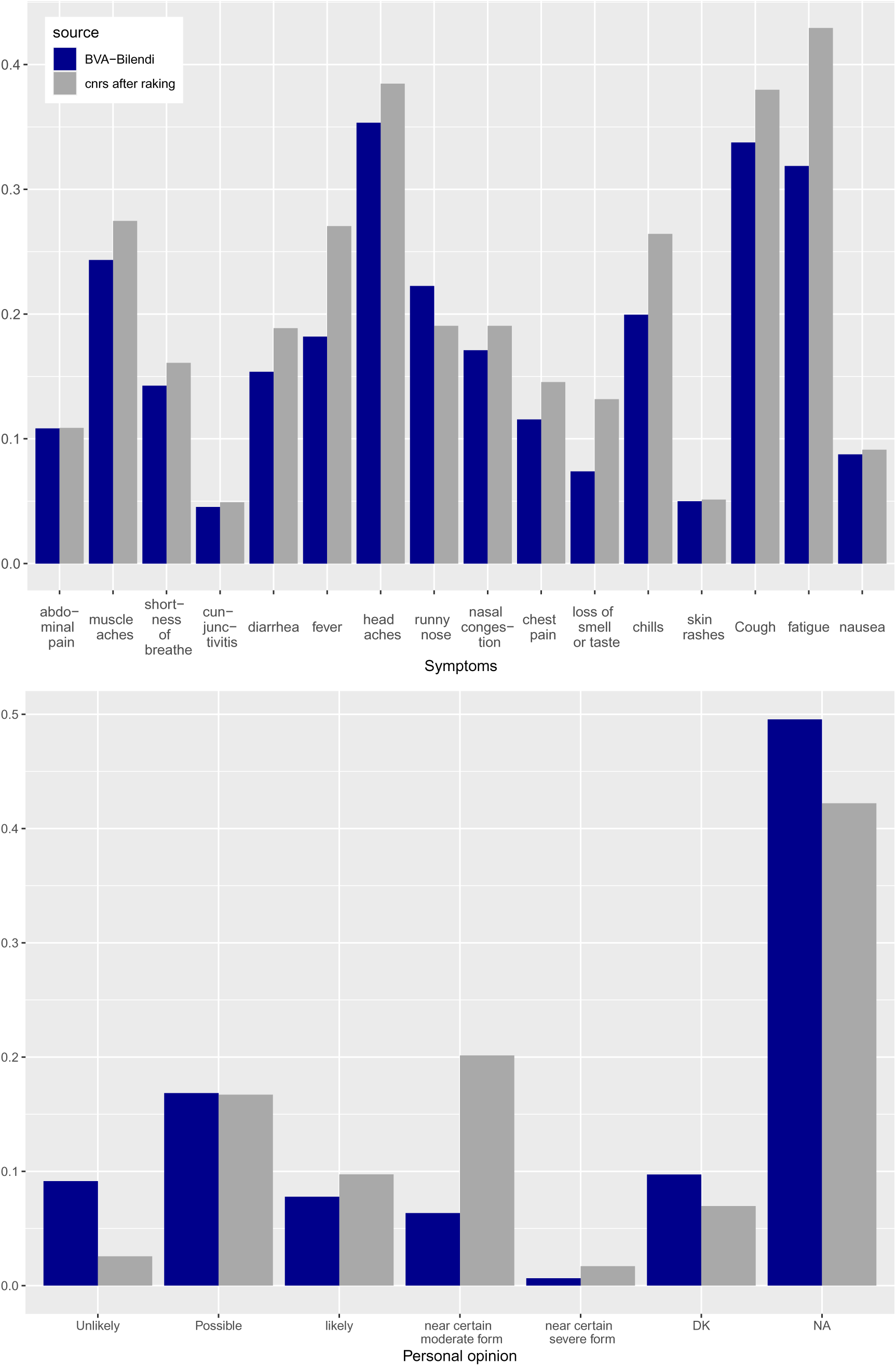
Proportion of individuals suffering from each symptom (top) and proportion of individuals with different personal opinions regarding the chances that they were infected (bottom). The results are displayed for the BVA-Bilendi dataset and for the CNRS dataset23after raking.

### 3.5 Which factors impact the presence of symptoms

We now consider a unique merged dataset, composed of the BVA-Bilendi dataset and the CNRS dataset weighted with the raking procedure described in Section 2.3.2. We investigated what features could be related to the presence of symptoms associated with Covid-19. In the following, we consider the binary variable ‘to have at least 1 symptom’ or not. We conducted the same analysis with the variable of interest ‘to have at least 2 symptoms’ or ‘to have at least 3 symptoms’, but the results are very similar. This is due to the fact that among people who presented symptoms, many had several of them and only a small group had only one or two symptoms, as we can see in Figure 8 (bottom panel). The most frequent symptoms are headache, cough and unusual fatigue (see Figure 8, top panel).

**Figure 8:**
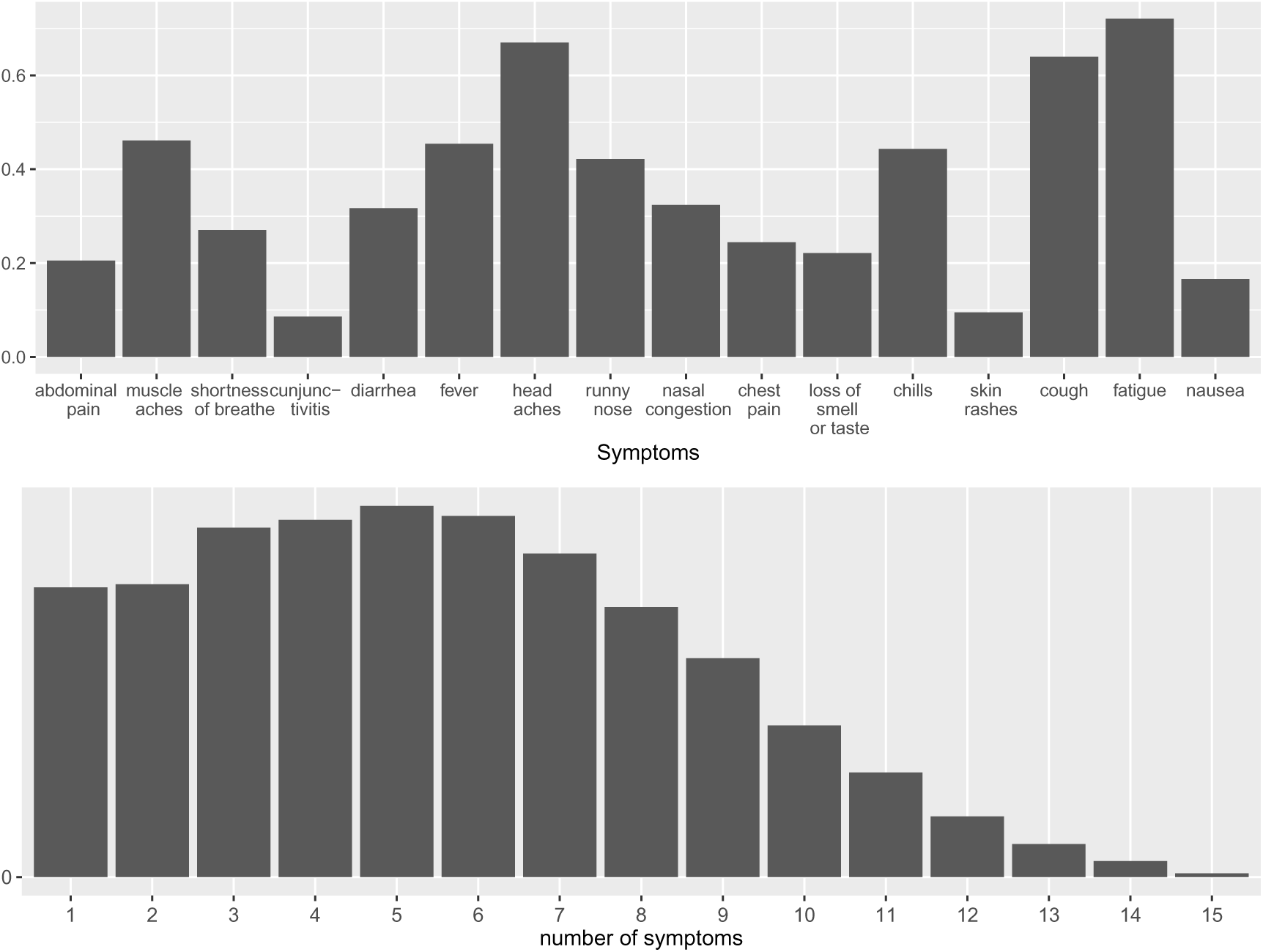
Frequency of each symptom (top) and distribution of number of symptoms (bottom) among the individuals who have at least one symptom (57.8% of the whole dataset).

We identify several factors associated with the presence of symptoms, such as weight and age. For example, overweight individuals show more symptoms, see Figure 9. The presence of symptoms increases with age up to 40 years, and then decreases with age. Symptoms are substantially rarer among children compared to any other age category. The age category that most expressed symptoms is adults between 25 and 40.

**Figure 9:**
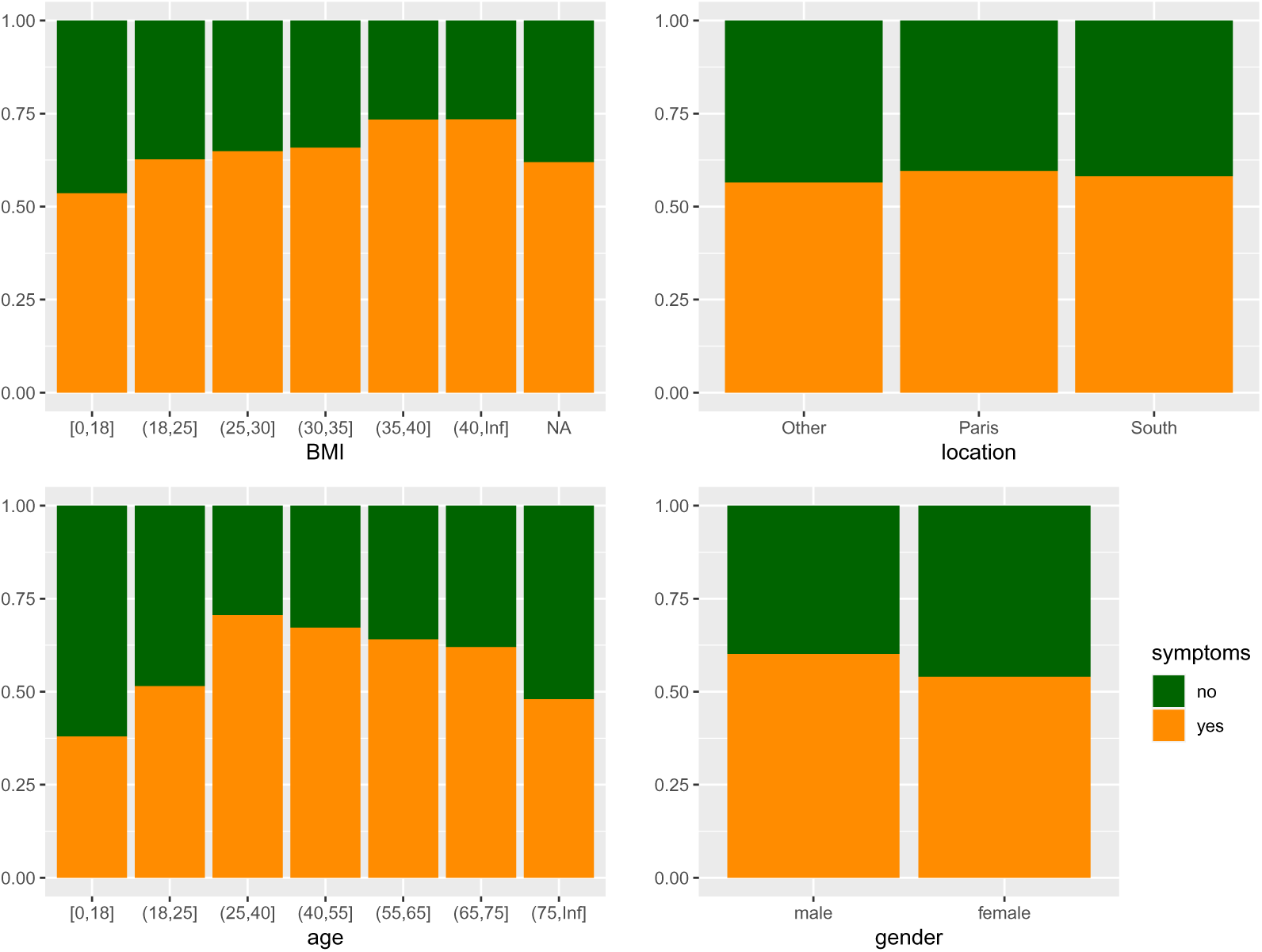
Proportion of individuals with/without symptoms among each BMI (top left), location (top right), age (bottom left), and gender (bottom right) categories.

The number of symptoms is also higher for male than female respondents and slightly higher in the Paris region than elsewhere in France.

We also focus on the influence of the spatial proximity of individuals within households, which is assessed according to the number of rooms and of people in the household. Figure 10 shows that people express less symptoms when their home contains more space and also when there are more people in the household, although both features are obviously linked. However, if we consider the ratio between the number of people and the number of rooms, which is a reasonable indicator of spatial proximity within the home, we see that the number of symptoms increases when there is more space and fewer people. To better understand this unexpected relationship between the number of people, the number of rooms, and the presence of symptoms, the heat map shown in Figure 10 shows that the presence of symptoms increases strongly in households that contain only two people. However, for a given number of people in the household, the presence of symptoms seems rarer in spacious households, which is expected. The interpretation of the impact of these different features will be discussed in Section 4.4.

**Figure 10:**
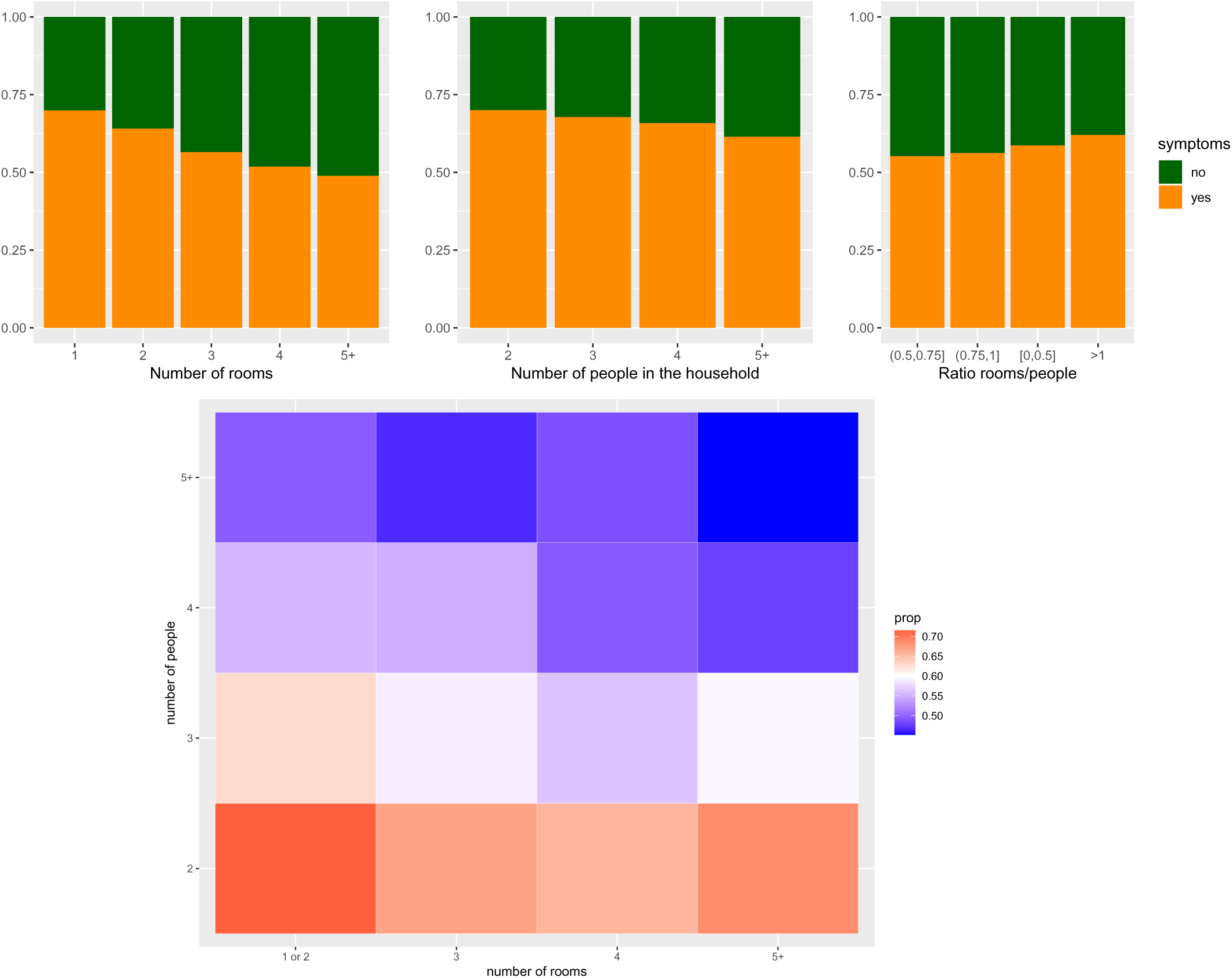
Top panel: proportion of individuals with/without symptoms depending on the number of rooms in the home (left), the number of people in the household (middle), and the ratio between the number of rooms and the number of people (right). Bottom panel: proportion of people with symptoms depending on both the number of rooms and the number of people in the household.

In order to assess the statistical significance of the results provided by the visual analysis, we perform a multivariate logistic regression with associated likelihood-ratio tests. We use the R library car to assess the significance of adding each feature to the model (the package performs the likelihood ratio test between the models with and without each feature). The very low p-values associated with the different features demonstrate their impact on the presence of symptoms, particularly for age, number of people in the household, and gender.

Table 5 displays the coefficients estimated using logistic regression for the model that contains the significant variables BMI, age, number of people, number of rooms, location, and gender. The larger coefficients correspond to the age categories that express more symptoms (adults between 25 and 65), the people with a strong overweight (BMI greater than 35). A strong negative coefficient is observed for large households (more than 4 people).

**Table 5:** Estimated coefficients for the logistic regression over BMI, age, number of rooms, number of people, gender, and location. The first column of each row corresponds to the reference category, the estimated coefficient of which is always equal to 0. The other coefficients are either positive (if the corresponding category increases the probability of having symptoms compared to the reference category) or negative (if the corresponding category decreases the probability of having symptoms compared to the reference category).

|  |  |  |  |  |  |  |  |
| --- | --- | --- | --- | --- | --- | --- | --- |
| Age | **[0, 18] (ref)** | (18,25] | (25,40] | (40,55] | (55,65] | (65,75] | (75,Inf] |
| Coeff | **0** | 0.062 | 0.22 | 0.20 | 0.14 | 0.096 | -0.04 |
| BMI | **[0, 18] (ref)** | (18,25] | (25,30] | (30,35] | (35,40] | (40,Inf] | - |
| Coeff | **0** | 0.050 | 0.068 | 0.070 | 0.14 | 0.12 | - |
| Nb people | **2 (ref)** | 3 | 4 | 5 | 6 | 7+ | - |
| Coeff | **0** | -0.063 | -0.096 | -0.11 | -0.099 | -0.18 | - |
| Nb rooms | **1 (ref)** | 2 | 3 | 4 | 5 | 6+ | - |
| Coeff | **0** | -0.0082 | -0.033 | -0.049 | -0.075 | -0.0071 | - |
| Gender | **Male (ref)** | Female | - | - | - | - | - |
| Coeff | **0** | -0.077 | - | - | - | - | - |
| Location | **Other (ref)** | Paris | South | - | - | - | - |
| Coeff | **0** | 0.034 | -0.017 | - | - | - | - |

The age is considered as a discrete variable with categories [0, 18], [18, 25], [25, 40], [40, 55], [55, 75] and more than 75. The BMI is considered as a discrete variable with categories [0, 18], [18, 25], [25, 30], [30, 40] and more than 40. The number of rooms is divided into 6 categories (from 1 to 5 and more than 5 rooms). The number of people is divided into 6 categories (from 2 to 6 and more than 6 people). The ratio between the number of people and the number of rooms is discretized in 3 categories: [0, 1], [1, 2] and more than 2. Gender is female or male. The location variable takes 3 values: Paris, South and other.

## 4 Discussion

### 4.1 Benefits and shortcomings of the protocol

In the beginning of an epidemic of an emerging disease, the temptation can be great to distribute the first diagnostic tests available on the largest scale possible to gain knowledge of its prevalence. In relation to the cost of this kind of initiative, the information it provides actually presents modest interest and will be outdated after a couple of months. In contrast, the household survey we described in this article is low-cost and provides information on both the sociodemography of the epidemic and the transmission mechanism within households. In addition, the design of the survey is easy to reproduce – future authors may wish to rely on the detailed example given in Section A.

We used data collected from two surveys: the CNRS open online survey and the representative BVA-Bilendi survey. Let us discuss the benefits and shortcomings of these two modes of administering questionnaires.

Because the online survey relies on self-reported cases, it suffers from numerous statistical biases, including the certain presence of symptoms and general biases due to snowball sampling (Rocha et al., 2017).

Our results show that the media campaign was essential to the diffusion of the survey: not only did it strongly increase the total number of respondents but it also allowed the increase of the diversity of the dataset, in particular in terms of geography (most early respondents lived in the largest cities in France). We recommend extending the period of the media campaign and increasing the time between interventions. First, the spread of responses over time can provide information on the dynamics of the epidemic. Second, the marginal benefit of an additional media intervention is greater when spaced in time. We also recommend diversifying the types of media (local vs national newspaper, TV channels, radio stations, etc.) to reach a wider variety of respondents in terms of social characteristics, although we were unable to investigate the impact of each type of media because interventions were often too close to each other (sometimes several on the same day).

Finally, we recommend paying attention to the simplicity of the URL to communicate, as journalists will not transmit anything complicated.

The BVA-Bilendi survey allowed us to obtain a sample which is representative of the French society. It gives a faithful snapshot of the state of the epidemic and its interaction with sociodemographic factors. This survey has been invaluable in allowing us to estimate the prevalence of the disease and correct for biases naturally present in the online survey. However, it is not always possible to count on the spontaneous contribution of survey institutes, as we had the luck to. An alternative option would be to use crowd-sourcing websites such as Prolific^3^ or SurveyMonkey Audience^4^ although these might also present biases.

An inherent difficulty of surveys is data privacy, accessibility, and storage. Respondents should be able to access their data, but the data should be pseudonymized. Data must be stored on secure servers, well protected from malicious access, but should be available to the research community. In reality, the fine balance is hard to find and data are either not accessible by anyone (respondents, research community) or accessible by virtually everyone who has hacking skills. Thus, the intervention of a data professional is strongly encouraged during questionnaire conception (to ensure proper pseudonymization), during data analysis (to ensure secure storage), and after publication (to ensure proper data sharing).

### 4.2 What we realized we should have put in the questionnaire

The questionnaire detailed in Section A is the result of a trade-off between the need for detailed information and the deterrent effect of long questionnaires on respondents. A specific effort has been made to disambiguate answers, for example regarding symptoms (fever *>*38°C, diarrhea ‘three soft stools in a row’, runny nose ‘except spring allergy’). The questionnaire could have been improved by further disambiguation, for example, the symptom ‘unusual fatigue’ can be made more concrete, as in ‘unusual difficulty to climb stairs’.

Had we had better knowledge of covid symptoms at the time, we would have added the loss of appetite as an additional symptom, concretely expressed, for example, as ‘skip two meals in a row’. The questionnaire could also have been improved by adding a question on the duration of symptoms, which we now know is a marker of infection by SARS-CoV-2 (Ashcroft et al., 2020) compared to respiratory infections with similar symptoms (influenza, common cold, respiratory syncytial virus…).

Finally, when implementing such an online questionnaire, it is important to give the user the possibility to correct past answers (‘back button’), to return later to the questionnaire (‘resume button’), and to signal that the questionnaire is complete (‘quit button’). Because we did not add a ‘quit button’, we had to discard some responses on the basis of the time taken to fill the questionnaire (too short, lower than 3 minutes, or too long, larger than 35 minutes).

Open-ended answers, especially quantitative ones, must be kept to a minimum due to possible errors, even when controlling for the format of each answer. In our case, we frequently received responses about size and weight that satisfied the required format but produced an unlikely high BMI. Thus, we recommend the discretization of variables into intervals of values that the user can choose or if not possible a posterior check of consistency between variables.

### 4.3 Comparison between the two populations after raking

We can see in Figure 5 that raking allowed us to efficiently correct for population differences when considering the very statistics used for correction (age, bmi, household size, etc.). In addition, raking indirectly corrected for differences in higher-level population characteristics, such as the numbers of households at risk or of parental households (see Figure 6). Some differences remain between the two populations even after raking, especially when considering the reported symptoms and the rate of confirmed covid (Figure 7). This is typically a bias due to the self-reported versus queried nature of the questionnaire in the two populations. Since the CNRS questionnaires have been self-reported by households who suspected at least one case of covid, this results mechanically in higher rates of reported symptoms and a much stronger opinion regarding the presence of covid cases.

### 4.4 Factors impacting symptoms

Among the features that we found associated with the presence of symptoms, some were expected and some were less obvious to interpret. The presence of symptoms that increased with BMI was expected (Gao et al., 2021). Our analysis also confirms the importance of age (Williamson et al., 2020) but infers a surprising effect of age on the expression of symptoms, namely, the age categories with more symptoms are adults between 25 and 65, and people over 75 show a very low proportion of symptoms. A possible explanation is that the behavior during lockdown was strongly dependent on age. Adults between 25 and 65 likely continued to leave their house on a regular basis, while older people tended to protect themselves and respect strict confinement. Another explanation is that elderly people with many symptoms may have previously been transported to the hospital.

Another difficult question is the effect of the density of people in the same household, as measured by the number of rooms divided by the number of people. We expected that density should increase the spatial proximity and the likelihood of transmission within the home, and hence the presence of symptoms (Cauchemez et al., 2009; Jing et al., 2020; Madewell et al., 2022). However, we rather observe the opposite effect. The heat map displayed in Figure 10 provides a possible explanation. Regardless of the number of rooms, households composed of two people generally express more symptoms: most of these households are likely couples who share the same bedroom. In contrast, large households with more than 5 people and/or 5 rooms, which are the category with the smallest proportion of symptoms, might include younger people, which are less likely to have symptoms (families, shared flats), with separate bedrooms. Therefore, the density of a household may not be sufficient to explain disease transmission within a home.

### 4.5 Conclusion

The methodology described here is a modernized version of an ancient procedure consisting of sampling households and counting the total number of people infected in each household. Considering these households as independent and identically distributed replicates of the same micro-epidemic, such data can help to estimate some parameters of the epidemic model, generally chosen to be the Reed-Frost model (Fraser et al., 2011). However, such a collecting procedure is only possible if symptoms are unambiguous or diagnosis tests are available. In addition, parameter estimation faces problems of identifiability (Jacquez, 1987) which can only be overcome by measuring finer data upon sampling, typically about symptom dates and durations, as we did here.

By having each household fill out a detailed questionnaire, we proposed and tested a tool refining the standard procedure with the aim of improving diagnosis and estimating more robustly parameters of the model of intra-household transmission. This tool can be quickly adapted and deployed for future outbreaks where overlapping symptoms and lack of reliable diagnosis tests are a problem. It can also be applied, for COVID-19 or any other infectious disease, in low-income countries with limited testing and surveillance facilities, to better understand the nature and dynamics of the outbreak. Alternatively, even if a massive release of diagnosis tests makes our tool obsolete, parameter estimates obtained early in the epidemic can prove useful to assess changes due, for example, to pathogen evolution, vaccination, seasonality or shifts in social behavior.

## Data Availability

All data produced in the present study are available upon reasonable request to the authors

## Acknowledgements

We are grateful to UMS GRICAD (Violaine Louvet) for their support in hosting, storing, and coding the CNRS online survey. We thank the numerous voluntary colleagues and friends who tested and disseminated the survey. We very warmly thank Bilendi (Marc Bidou, Stéphanie Sanchez Incera) and BVA (Odile Peixoto, Elodie Laville) for their pro bono assistance in recoding and administering the survey to their representative panel, which was essential for data weighting. We also acknowledge Isabelle Hilali (datacraft) and Xavier Fresquet (Sorbonne cluster for artificial intelligence) for their help and for connecting us with Ekimetrics (Soline Aubry), who generously provided pro bono support through data storage and analysis. We thank the Modcov19 Task Force (Emmanuel Royer, Jean-Stéphane Dhersin, Pétronille Danchin) for their advice and connections with the media, as well as the communication services of Sorbonne Université (Claire de Thoisy-Méchin, Marion Valzy) and of Collège de France (Raynald Belay, Guillaume Kasperski). We are grateful to the Scientific Advisory Board of Alcov2 (Jean-Christophe Thalabard, Vincent Maréchal, Odile Launay, Marie Jauffret-Roustide, Isabelle Hilali) for their continuous guidance regarding infectious diseases, survey design, and statistical methodology. All of these contributions were achieved in record time, driven by a genuine commitment to serving society.

## Authors contributions

- Amaury Lambert: Project conceptualization, Methodology (survey), Methodology (model), Project administration, Supervision, Writing.
- Anna Bonnet: Methodology (data analysis), Data analysis (visualization and statistical analysis), Writing.
- Pierre Clavier: Data analysis.
- Pierre Biousse: Data analysis.
- Lucas Clavières: Data analysis.
- Sophie Brouillet: Methodology (survey), Coding (survey).
- Sylvie Chachay: Methodology (survey), Coding (survey).
- Marie Jauffret-Roustide: Methodology (survey), Methodology (data analysis).
- Sonia Lewycka: Methodology (survey).
- Nicolas Chesneau: Methodology (data analysis), Data analysis, Project administration, Supervision.
- Grégory Nuel: Methodology (survey), Methodology (model), Methodology (data analysis), Data analysis, Project administration, Supervision, Writing.

## A Questionnaire

### A.1 Questions only included in the BVA/Bilendi survey

In the BVA-Bilendi survey, the questionnaire first asked zip code and name of municipality of usual residence place, hence recording department, region^5^ and agglomeration size, for quota adjustment.

Then the respondent was asked if h/she is the reference person in the household (‘personne de référence’, i.e., the active person who brings in the most income) and what is the profession of respondent/of reference person (if different), with the following choices:

1. Farmer (self-employed)
2. Craftsman, merchant, small business owner
3. Business owner
4. Liberal profession (except paramedical)
5. High school teacher, scientific profession
6. Executive and other higher intellectual profession
7. Foreman, supervisor, paramedical profession, technician
8. Schoolteacher or equivalent
9. Clerk
10. Maintenance staff
11. Worker, agricultural worker
12. Retired
13. Pupil, student
14. Other inactive (at home, in some other situation)

Last, the respondent is asked to give the zip code of her/his confinement residence place.

### A.2 Questions common to both surveys

The survey is entitled:

‘Alcov2 – An anonymous questionnaire to study the presence of symptoms possibly related to Covid-19 and the history of their transmission in French households during the lockdown period.’

Then the questionnaire starts with the following introductory text:

“This survey was set up by researchers from Sorbonne Université, Collège de France and Oxford University. Its objective is to study how the new SARS-CoV-2 coronavirus is transmitted within the same confinement residence place and the role played by asymptomatic or pauci-symptomatic individuals (carriers of the virus but showing no or few symptoms) in this transmission.

Information on the dates of symptom onset in different members of the confinement place is important because it can provide researchers with information on the incubation period of the disease, the mode and speed of transmission of the virus within the same outbreak, and the possible presence of infected but asymptomatic individuals.

These data, in combination with seroprevalence studies, will allow an understanding of how the virus was transmitted during the lockdown period, estimate the number of immune individuals who had little or no illness, and predict the effect of targeted quarantine measures after the end of lockdown.

Thank you for completing this survey:

- If at least one person living in your confinement residence place has experienced one or more symptoms, even mild, since mid-February that could be related to Covid-19, even if you have no idea if they are really due to Covid-19, for example: fever, cough, headache, sore throat, fatigue, diarrhea, muscle aches, chills, nausea, loss of taste or smell, shortness of breath, chest pain.
- If there are at least two (i.e., two or more) people in your confinement residence place.” We now reproduce the questions and their possible multiple answers.

**Part I. Questions about residence place.** The questionnaire starts with the following short questions on the location, size and constitution of household.

1. In which department is located your confinement residence place?
2. How many people live in your confinement residence place (including yourself and children even in shared custody)? [Answer hereafter denoted *X*]
3. How many bedrooms are there in this place?
4. Thank you for listing the ages of all *X* people living in this place.

[The algorithm then allocates a denomination to each person, for example: “Adult 2 (42 years old)”, depending on age, according to the following classification: Child (0-11), Teenager (12-17), Adult (18-64), Senior (*>*64).]

**Part II. Medico-social characteristics of household members.** The following list of questions unfolds for each of the *X* persons sharing the residence. Phrases in brackets were visible to the respondent. Multiple proposed answers are indicated between square brackets and ‘DK’ stands for ‘Don’t Know’ (‘Ne se prononce pas’ or ‘Ne sait pas’ in French). Apart from those, everything in square brackets was invisible to the respondent.

1. What is the biological sex of this person? [Check: Male/Female]
2. [If age *>* 2 years] What is approximately the size (cm)/weight (kg) of this person?
3. Has this person been absent from the residence for at least 3 consecutive days and nights since beginning of lockdown? [Check: Yes/No/DK]
4. Is this person absent from the residence at least 3 days a week because of her/his work or activity [Check: Yes/No/DK]?
5. [If Yes] for this job or activity, is this person in contact with the public? [Check: No/Yes, hospital or medical facility/Yes, other (sales, public transport, police…)/DK]
6. Is this person vaccinated against tuberculosis (BCG, which leaves a small scar on the upper left arm)? [Check: Yes/No/DK]
7. [If adult woman] Has this person been pregnant for more then 3 months? [Check: Yes/No/DK]
8. Does this person smoke or has this person smoked regularly (smokes/vapes at least once a day)? [Check: Yes, now/Yes, in the last 3 years/Yes, in the last 10 years/Never/Other]
9. Does this person suffer from a chronic disease? [Check: Yes/No/DK, if Yes, check: Diabetes/Respiratory disease/Heart or vascular disease/Kidney disease/Liver disease/Immuno suppressive disease or treatment/Other]
10. Since mid-February, has this person experienced one or more symptoms, even mild, possibly related to Covid-19? (for example: fever, cough, headache, sore throat, fatigue, diarrhea, chills, nausea, loss of taste or smell, shortness of breath, chest pain, etc.)? [Check: Yes/No/DK]

[The algorithm keeps in memory the identity of each of the *Y* persons having experienced symptoms (answer ‘Yes’ to the previous question). If *Y* = 0, the survey stops here.]

**Part III. Date of appearance of symptoms for each putative case.** The respondent is asked to rank chronologically, by date of symptom onset, the *Y* persons having experienced symptoms, hereafter referred to as ‘cases’. Then the respondent is asked to be more specific and give a date of symptom onset for each case, or to give the approximate durations elapsed between symptom onsets of each pair of consecutive cases.

**Part IV. Date of appearance and list of symptoms for each putative case.** The following list of questions unfolds for each case, in the chronological order completed in Part III. Multiple proposed answers are indicated between square brackets and ‘DK’ stands for ‘Don’t Know’ (‘Ne se prononce pas’ in French).

1. Which of the following symptoms did this person experience? [Check: Yes/No/DK for each item]

- Fever (*>*38°C)
- Chills
- Unusual fatigue
- Headaches
- Loss of smell or taste
- Muscle aches
- Cough, sore throat
- Shortness of breath
- Chest pain or tightness
- Abdominal pain
- Nausea, vomiting
- Diarrhea (three soft stools in a row)
- Nasal congestion
- Runny nose (except spring allergy)
- Conjunctivitis
- Skin rashes

2. Did these symptoms require this person to seek medical attention? [Check: Yes/No/DK]
3. [If Yes] What was the doctor’s diagnosis? [Check: Covid-19 unlikely/possible/likely/certain or near-certain/DK]
4. Has this person had a diagnosis test for Covid-19 (nasopharyngeal swab/ oropharyngeal swab/blood sample)? [Check: Yes/No/DK]
5. [If Yes] What was the result of this test? [Check: Positive (infection)/Negative (no infection found)/DK]
6. Has this person been hospitalized because of her/his symptoms? [Check: Yes/No/DK]
7. And you, do you think this person was sick with Covid-19? [Check: Covid-19 unlikely/possible/likely/near-certain, moderate form/near-certain, severe form/DK]

## Footnotes

1 see below for the accurate interpretation of ‘recently’

2 The administrative geography of France is divided into 99 *départements* (hereafter ‘department’).

3 https://www.prolific.com –Accessed: 2025-09-16

4 https://www.surveymonkey.com/mp/audience –Accessed: 2025-09-16

5 The administrative geography of France is divided into 12 ‘*régions*’ (or ‘region’) and 99 *départements* (or ‘department’).

